# COVID-19 Active Surveillance Simulation Case Study - Health and Economic Impacts of Active Surveillance in a School Environment

**DOI:** 10.1101/2020.10.28.20221416

**Authors:** Ali A. Saad, Malak Saad, Emad M. Boctor

## Abstract

The COVID-19 pandemic has affected the lives of almost all human beings and has forced stay-at-home mandates across the world. Government and school officials are facing challenging decisions on how to start the new 2020/2021 school year. Almost every school system has chosen a remote learning model for the Fall of 2020 while many are facing financial and logistical challenges.

In this study, we explore the efficacy of an Active Surveillance testing model where a random number of students are tested daily for early detection of asymptomatic patients and for prevention of the infection among the student population. In addition to health impacts, we also analyze the financial impact of deploying the Active Surveillance system in schools while taking into consideration lost workdays of parents, hospitalization costs, and testing costs.

Under the given assumptions, initial modeling results indicate that low Active Surveillance testing rates (between 6-10% daily testing of student population) can help achieve low infection rates (≤10%) among students along with enforcing mitigation procedures, such as wearing masks and social distancing. Without enforcing mitigation procedures, the optimal Active Surveillance rate of 8-10% can also achieve (≤10%) infection rates among student population. The results also demonstrate that Active Surveillance can lower the financial burden of the pandemic by proactively lowering the infection rates among student populations.

## Introduction

According to the Johns Hopkins Coronavirus Resource Center^**1**^ as of October 2020, global cases of COVID-19 pandemic exceeded 44 million people while death toll exceeded 1.7 million people. Many countries have issued stay-at-home or lock down orders to control the spread of the virus. The vast majority of schools around the world have been cancelled since March 2020 and up to this date the school system is struggling on how to start the new academic year of 2020/2021. Many school systems have chosen a virtual learning model while a few have chosen a hybrid system of in-person and virtual learning while enforcing social distancing and mask wearing mandates. Education and school officials try to make hard decisions to balance the health and safety of students, their families, their teachers, and the quality/efficacy of face-to-face education.

On the positive front, diagnostic tests for COVID-19 are now readily available. PCR-based tests can diagnose active patients. Meanwhile, Serology tests are blood-based tests that can detect COVID-19 antibodies. Test times can range from minutes to hours to even days.

In this paper we study an Active Surveillance Model for proactively detecting COVID-19 infections among school students. Our Active Surveillance model involves random daily testing for a percentage of students for early detection and quarantine of sick students. We study the impact of Active Surveillance on infection rates and health of our students as well as the economic impacts of reducing quarantine and hospitalization rates.

## Simulation Study Design

There have been a few publications on simulating the spread of the COVID-19 among populations, the most famous of which is the Washington Post article^**2**^ involving the simulation of COVID-19’s exponential spread and several suggested ways to flatten the curve using social distancing and mask wearing practices.

We based our simulation model on the Coronavirus Simulation Matlab program written by Joshua Gafford^**3**^, which is a recreation of the Washington Post COVID-19 simulation article listed above. The Matlab code simulates COVID-19 transmission among a human population in a confined space. A simple multibody physics model, elastic collision between two equal-mass particles, is applied to determine people’s trajectory within the space.

We modified the “Stimulus” Matlab code to represent a typical school environment where students and teachers interact together on a daily basis. Our school population was 500 people and the simulation’s assessed a 60-day duration to represent a school quarter. Some critical simulation parameters include: the probability of carriers initially infected (fixed at 1%), the probability of disease transmission (99% for the normal behavior without any mitigation practices, such as social distancing and mask wearing, 30% or 60% for mitigation practices), mortality rate (fixed at 0.69% for the student young age group), average recovery period for sick students (14 days.) We assumed the worst case of asymptomatic disease pattern where sick students would still show up to school and possibly infect others.

The simulation tracks four metrics during the 60-day period: percentage of unaffected students, who did not catch the virus, percentage of infected students, percentage of recovered students from infection, and percentage of potentially deceased students based on the mortality rate. The three main scenarios for simulations are: 1) normal behavior with no mitigation practices, 2) mitigation practices inside the school including social distancing and mask wearing, and 3) active surveillance procedures where a percentage of students are randomly tested on a daily basis to detect and quarantine infected students before they infect others. The combination of Active Surveillance and either normal behavior or mitigation practices is also modeled.

## Simulation Results

The following set of figures will demonstrate some of the simulation results under different scenarios. Figure 1 demonstrates the baseline scenario of “Normal Behavior” without any mitigation practices (no mask wearing and no social distancing.) The left side shows a simplified simulated school environment where students, and teachers, move and interact daily. The color-coded dots represent students with different states (green: unaffected, red: infected, blue: recovered from infection, and black: deceased from infection.) The graph on the right side demonstrates the progression of the four metrics (unaffected%, infected%, Recovered%, and Deceased%) during the 60-day period.

**Figure 1.**
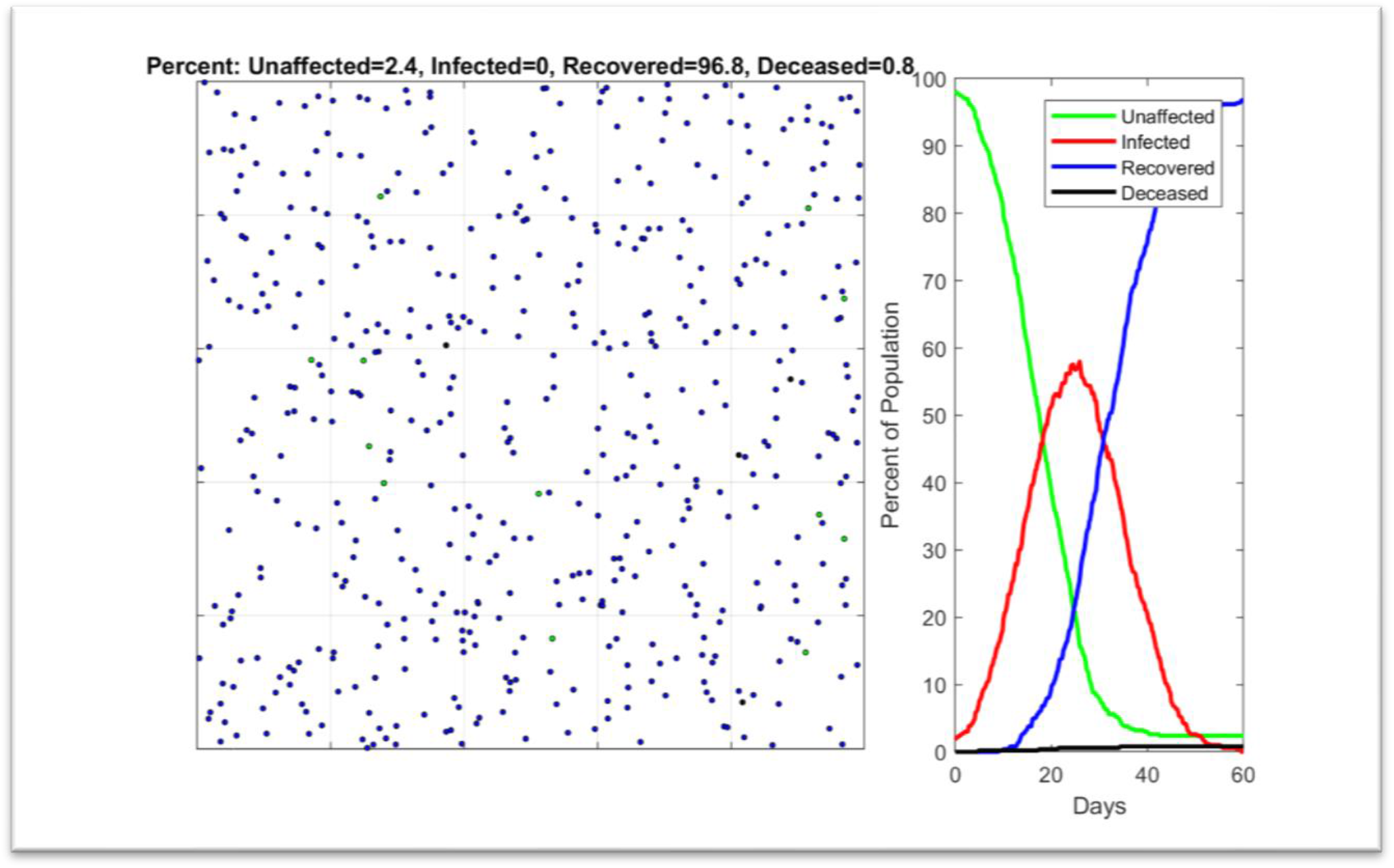
Simulation result for Normal Behavior. Left: simulation of student population. Right: simulation metrics: unaffected%, Infected%, Recovered%, and Deceased%. Infection rate is 97.6% (2.4% unaffected rate)

For the baseline normal behavior simulation case, the majority of the students (97.6%) will get infected by the end of the 60-day period as demonstrated by the 2.4% unaffected rate on Day 60. However, by the end of the period, 96.8% of students would recover after 14-day quarantine on average, while 0.8% of students could unfortunately pass away. Some of the infected students could potentially need hospitalization. We can conclude from this simulation case that it will be extremely difficult to return schools to normal practice if there are no mitigation behaviors implemented.

Figure 2 shows the simulation case of applying mitigation practices among students, such as mask wearing and social distancing. In this case we assumed the mitigation practices will decrease the disease transmission rate from 99% to 30%. We assume the mitigation practices will be enforced by school administrations to achieve such a high efficacy for decreasing the transmission rate. In this case the percentage of unaffected students drastically increased from the previous baseline case to 87.6% with only 12.4% infection rate) among the student population. This case proves the efficiency of the mitigation practices if strictly adhered to, however the challenge is how to enforce it among students especially at younger ages in elementary grades for example. To show how relaxing these mitigation practices can affect the outcome, we changed the disease transmission rate from 30% to 60% to simulate partial adherence to face masking and social distancing rules. Figure 3 shows the result of this case and the unaffected % has drastically decreased from 87.6% to 13.6% (86.4% infection rate)

**Figure 2.**
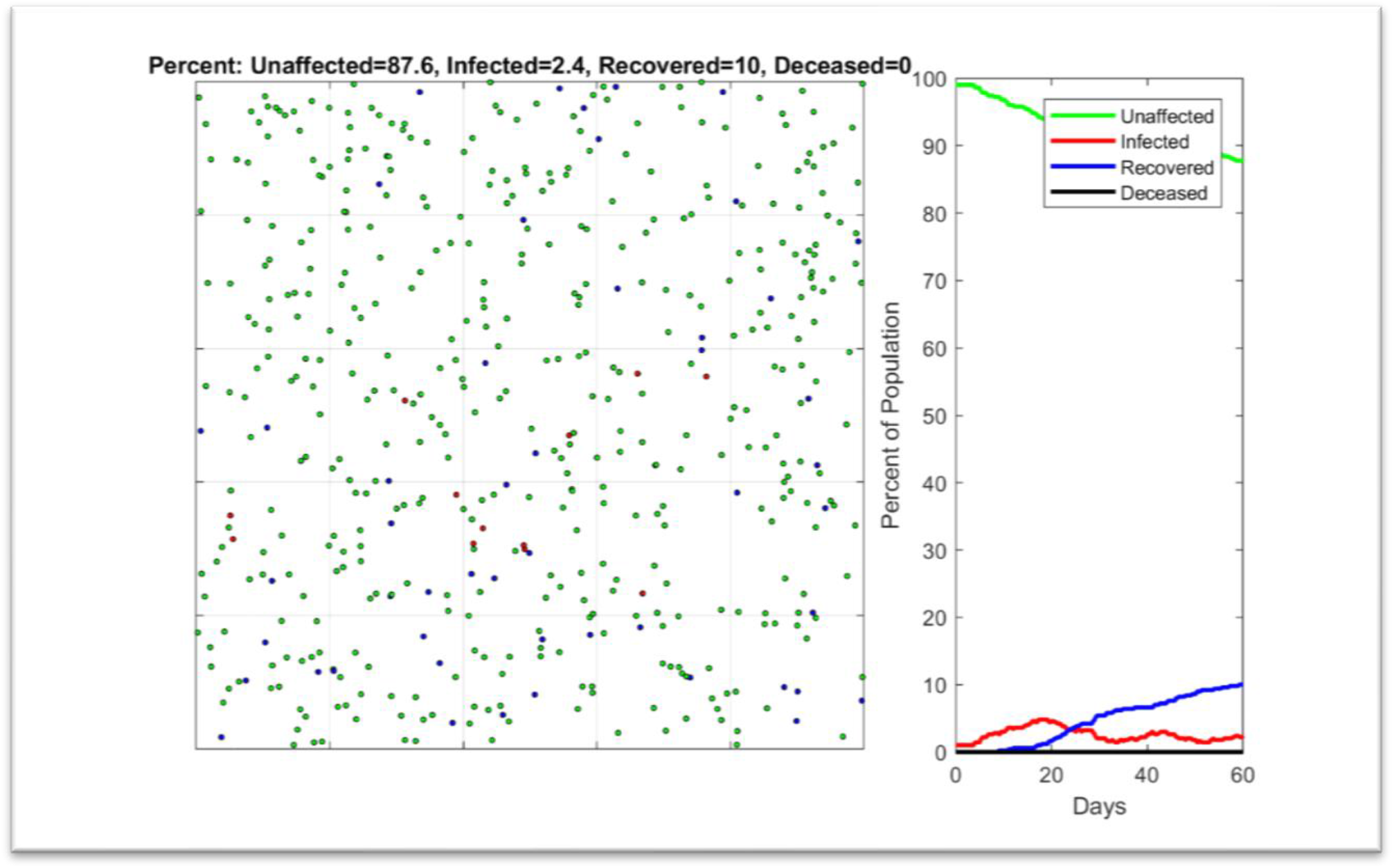
Simulation result for Mitigation Behavior (social distancing and mask wearing) with 30% infection transmission rate. Left: simulation of student population. Right: simulation metrics: unaffected%, Infected%, Recovered%, and Deceased%. Infection rate is 12.4% (87.6% unaffected rate)

**Figure 3.**
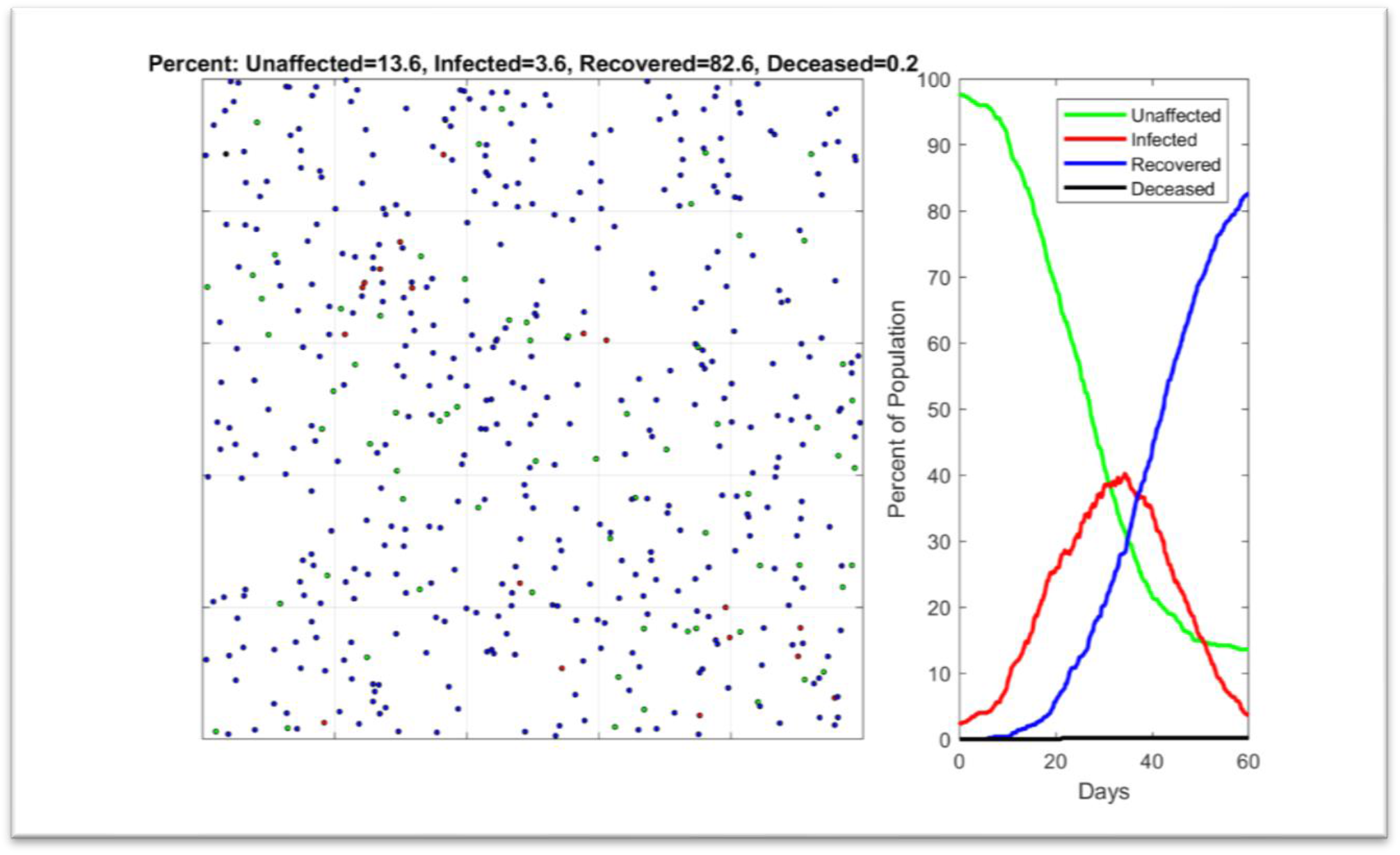
Simulation result for Mitigation Behavior (social distancing and mask wearing) with 60% infection transmission rate. Left: simulation of student population. Right: simulation metrics: unaffected%, Infected%, Recovered%, and Deceased%. Infection rate is 85.2% (14.8% unaffected rate)

To study the impact of Active Surveillance for COVID-19 testing, we designed a model to test a randomly selected percentage of students daily. For simplicity we assumed 100% test accuracy and we assumed immediate test results available to detect infected students before they can interact with their peers. Figure 4 demonstrates an Active Surveillance case of testing only 5% of the student population (25 tests/day) along with applying social distancing and mask wearing practices, which we assumed similar to Figure 3 of 60% disease transmission probability. The infection rate has drastically improved from 86.4% (13.6% unaffected rate) in Figure 3 to 12.8% infection rate (87.2% unaffected rate) in Figure 4. This result clearly demonstrates the efficacy of the Active Surveillance methodology even with a reasonable test rate (5% of students)

**Figure 4.**
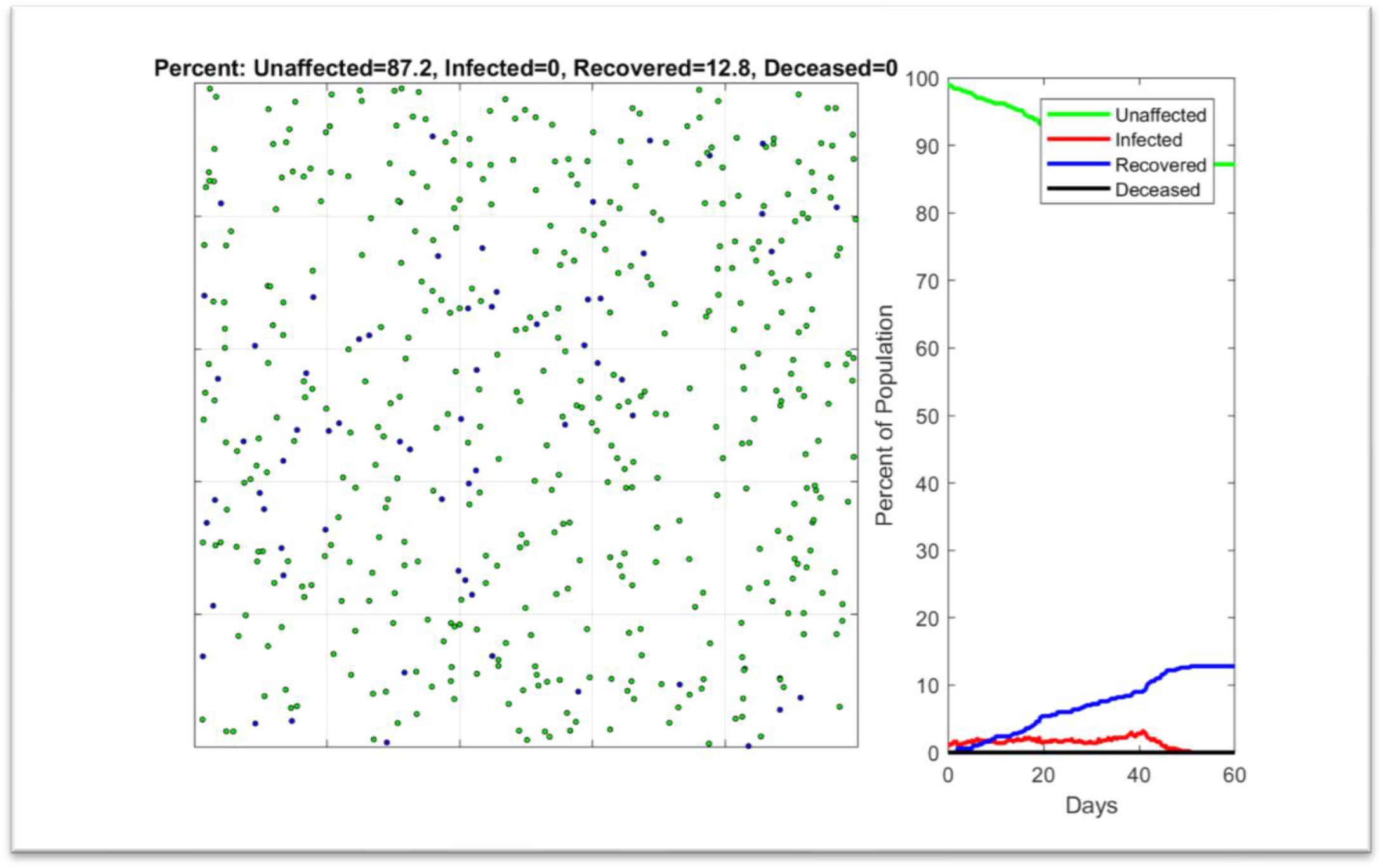
Simulation result for 5% Active Surveillance rate with Mitigation Behavior (social distancing and mask wearing) with 60% infection transmission rate. Left: simulation of student population. Right: simulation metrics: unaffected%, Infected%, Recovered%, and Deceased%. Infection rate is 12.8% (87.2% unaffected rate)

To test the sensitivity of the Active Surveillance approach, we lowered the daily test rate to only 1% of students (5 students out of 500.) Figure 5 shows the result of this scenario and it shows the infection rate increased to 74.4% (25.6% unaffected rate), which implies the non-efficient result if the Active Surveillance rate is too small.

**Figure 5.**
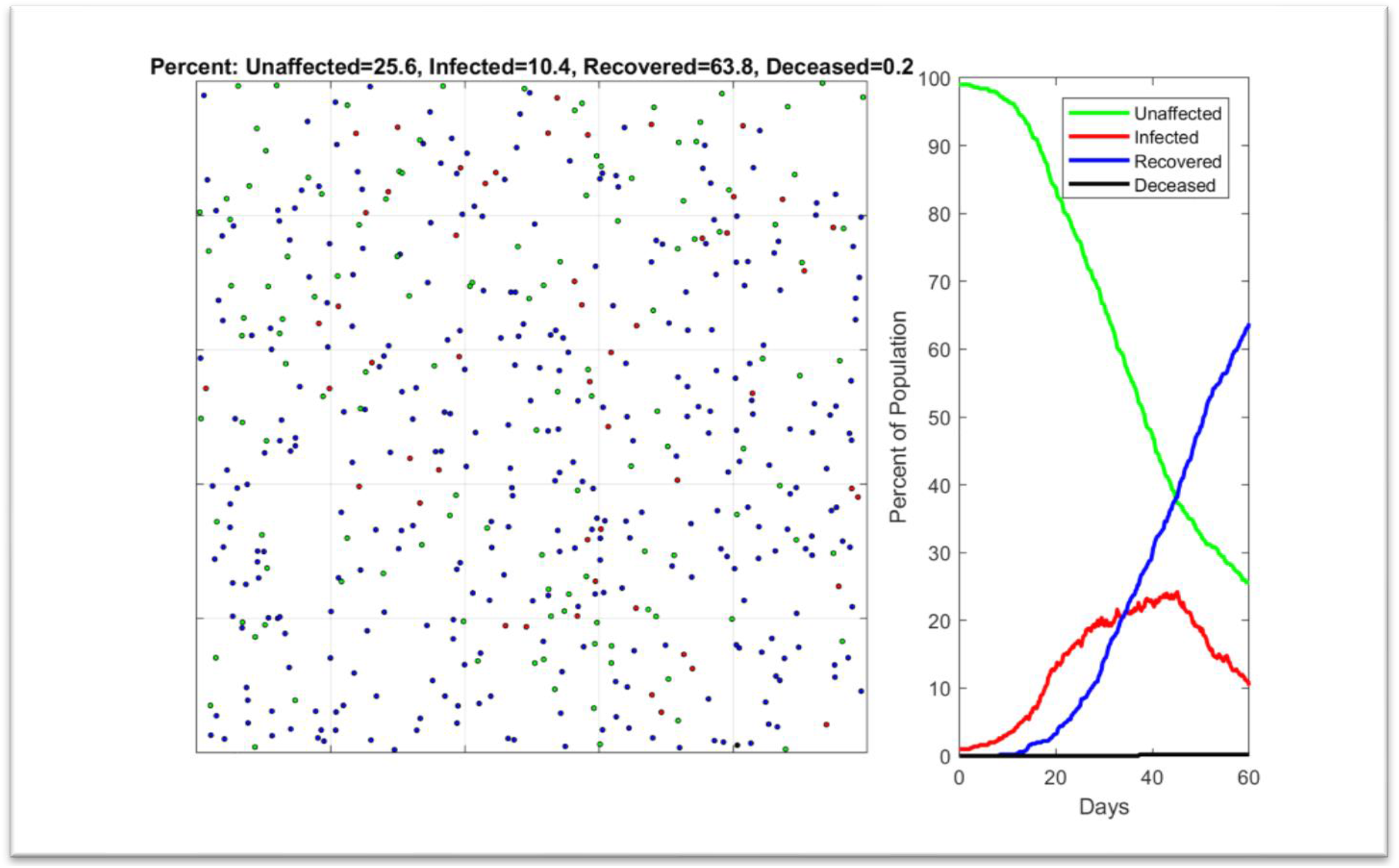
Simulation result for 1% Active Surveillance rate with Mitigation Behavior (social distancing and mask wearing) with 60% infection transmission rate. Left: simulation of student population. Right: simulation metrics: unaffected%, Infected%, Recovered%, and Deceased%. Infection rate is 74.4% (25.6% unaffected rate)

### Active Surveillance Health and Economic Impact

The previous simulations demonstrated the health benefits of applying Active Surveillance measures by reducing the infection rates among students. However, the economic impact of Active Surveillance is also an important factor when it comes to the financial burden of testing. The main question we try to answer in this section is, will the Active Surveillance methodology improve or disprove the financial burden of the COVID-19 pandemic?

Incorporating all the financial factors related to COVID-19 is a complex issue. In our study we incorporated three main cost metrics: 1) The cost of the daily Active Surveillance test, 2) The cost of potential hospitalization of some infected students,3)The cost of lost income of one or two parents who may have to take a leave at work to care for their sick student.

We tried to make some reasonable assumptions for the different cost metrics as follows. We assumed an average test cost of $50, a hospitalization rate of 5% among the infected population, an average hospitalization cost of $40K, and two weeks of lost income of $2,430, assuming that the 2019 U.S. median household income was $63K.

We ran our simulations while sweeping the Active Surveillance percentage from 1% to 100% with non-uniform intervals. In each run we average 20 cycles with the same settings and calculate the average infection and unaffected rate for students. We also calculate the daily test cost, daily potential hospitalization/income loss cost, daily total cost of testing, hospitalization, and income loss. Our goal is to find the optimal Active Surveillance test rate that can maximize the health benefits (lower infection rates) while minimizing average cost associated with COVID-19.

Figure 6 demonstrates how the unaffected rate (and thus the infection rate) and the average daily cost change as the Active Surveillance rate changes.

**Figure 6.**
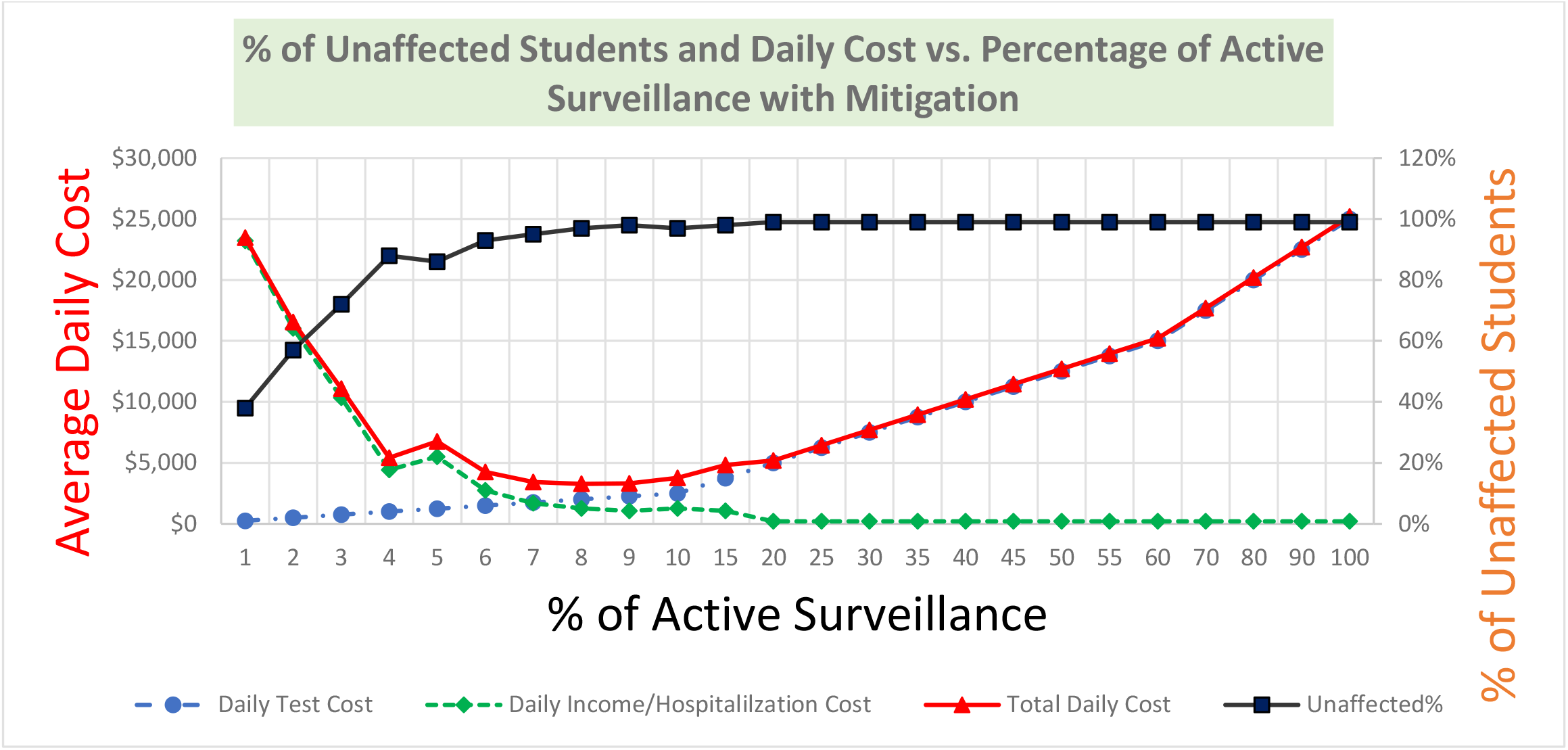
Active Surveillance % vs. Unaffected Student % vs. Average Daily Cost for the case of mitigation procedures with 60% disease transmission rate.

As the Active Surveillance test rate increases the unaffected rate increases (infection rate decreases) while the daily cost decreases due to minimizing hospitalization and income loss costs. After a certain Active Surveillance rate, around 6-10%, the infection rate becomes stable with no improvement of increased test percentages, while the daily average cost increases monotonically as more tests are being conducted daily. It is evident from the graph that the optimal test percentage could be anywhere between 6-10% of students getting tested every day to achieve the optimal minimal infection rate (≤10%)) and minimal average cost as well.

To evaluate the efficacy of the Active Surveillance testing alone without any mitigation procedures, we ran our simulations with no mitigation actions, no social distancing and no mask wearing where infection transmission rate is 99%. Figure 7 shows the results of normal behavior while sweeping the Active Surveillance test rate.

**Figure 7.**
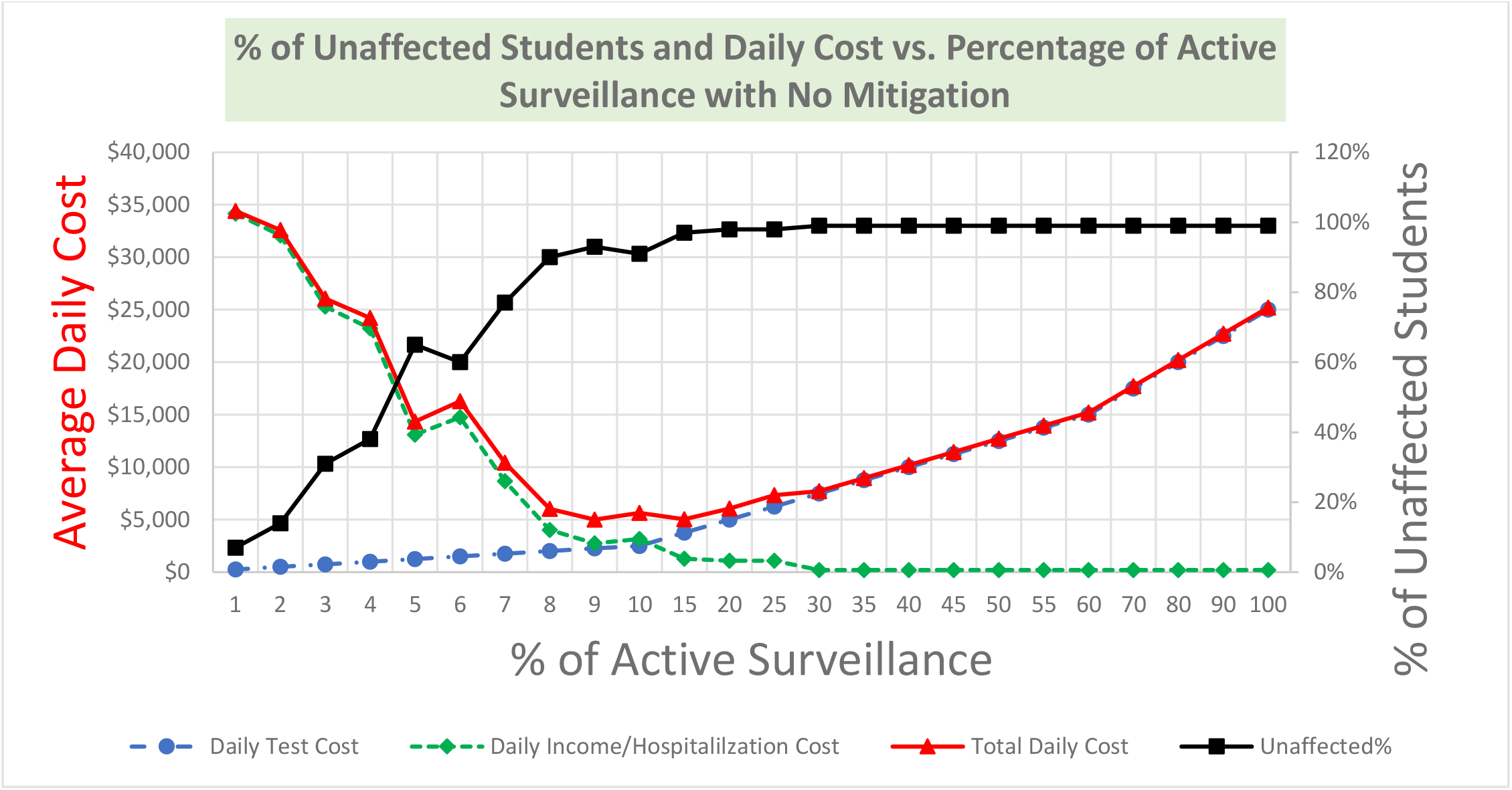
Active Surveillance % vs. Unaffected Student % vs. Average Daily Cost for the case of normal behavior with no mitigation procedures at all.

The unaffected rate graph and cost graph follow the same pattern as with the mitigation procedures but with pushing the optimal testing rate to 8-10% that can achieve the lowest infection rates (≤10%) and lowest average cost. This case may be useful to the elementary school student populations where enforcing the mask wearing and social distancing protocols may cause challenges.

## Conclusions and Future Work

In this work we investigated a simulation model for a school environment to better understand the impacts of social distancing, mask-wearing, and Active Surveillance testing on the infection rates among students. It is evident that the mitigation measures and the Active Surveillance testing are both effective in limiting the spread of the COVID-19 infection among students. The most effective practice is to employ both mitigation and Active Surveillance testing together.

Under the given assumptions, Active Surveillance with a reasonable test percentage (6%-10% with social distancing and mask wearing, or 8-10% without mitigation procedures) can achieve both health and economic goals of safe schools with lower economic burdens. No compromise needs to be taken to optimize both the health of our students and teachers and the economic burden of COVID-19.

For future work, we plan to incorporate more realistic assumptions about the speed of the test results, different test techniques and pricing, pool testing techniques (testing multiple samples with one test kit), student grouping (classroom divisions), etc. We will also investigate the sensitivity of the results with varying different parameters in our study, such as varying the cost of testing, test sensitivity and specificity, etc.

We hope this work helps inform and inspire our education and government leaders to consider Active Surveillance testing as a venue to reopen our schools safely and efficiently.

## Data Availability

N/A

